# Measuring Intertrochanteric Fracture Stability with a Novel Strain-Sensing Sliding Hip Screw

**DOI:** 10.1101/2020.09.04.20183251

**Authors:** Nathan T. Carrington, Paul W. Milhouse, Caleb J. Behrend, Tom B. Pace, Jeffrey N. Anker, John D. DesJardins

## Abstract

**Background:** Bone healing after internal fixation of intertrochanteric hip fractures is difficult to monitor with radiography, particularly with internal fixation implants such as the sliding hip screw (SHS). In this study, we evaluate a robust, user-friendly device to non-invasively determine the loading on the screw implant. This will allow clinicians to better monitor the status of bone healing and take preventative steps if complications occur.

**Methods:** A novel strain-sensing sliding hip screw (SS-SHS) was designed and refined using a finite element model of a simple intertrochanteric fracture and a standard SHS implant. The SS-SHS houses an internally fixed indicator rod, whose position relative to the screw body can be viewed on plain film radiographs to measure screw bending. Screw bending was assessed in an intact femur and an unstable A1 intertrochanteric fracture using a finite element computational model and compared with experimental axial loading of a femoral Sawbones composite and human cadaveric femur specimens. Indicator rod position relative to the screw was visually tracked using plain radiographs at each load state.

**Results:** The indicator rod was found to displace linearly in response to implant strain in the unstable fracture. This movement was consistently visible and measurable using radiography throughout loading cycles across the mechanical and cadaveric fracture models. Sensor movement was not detected in healed fracture models. The slope of the curve was approximately equal in the computations, composite and cadaveric models (0.08 µm/N, 1.0 µm/N, 0.08 µm/N, respectively). The noise level was approximately 25 N in the composite model and 63 N in the cadaveric specimen and this was sufficient to see 1/10^th^ of body weight or more for an 80 kg patient which is likely good enough to track fracture healing.

**Conclusions:** In current practice, clinicians must carefully monitor their patients for signs of implant failure after surgery. However, by the time signs of failure are apparent, it is often too late to avoid revision surgery. This device enables clinicians to quantitatively track fracture healing, and better communicate the process to the patient. Clinicians can also take preventive measures with at-risk patients before revision surgery is needed, thus reducing mortality risks.

**Clinical Relevance:** By augmenting an existing SHS system with an indicator rod, crucial information on the status of fracture healing can be ascertained from follow-up radiographs already taken with no additional risk to the patient.

## Introduction

Hip fractures are a common but significant injury for elderly patients, with over 330,000 annual occurrences in the United States alone.^1^ Approximately half of these fractures are intertrochanteric (IT) and are often treated with internal fixation implants such as the sliding hip screw (SHS).^2^ This approach allows patients to maintain their natural hip structure and regain functional mobility, and has long been preferred by surgeons for allowing deep implant insertion and controlled collapse of the fracture site.^3-5^ However, when treating these patients, surgeons must take special care in monitoring the facture site for adequate postoperative healing and take appropriate action if delayed fracture healing occurs.^6^ Failure to recognize a nonunion and take appropriate interventional measures could result in catastrophic fixation failure and the need for emergency revision surgery.^7,8^ Hip fracture patients are often frail and thus these revision surgeries are risky for them: the 1-year all-cause mortality rate for patients readmitted within 30 days of surgery is 56%, compared to 19% for those not readmitted.^9^ The average hip fixation hardware failure rate is around 5%; however, this can increase to over 20% in patients with comorbidities such as osteoporosis, unstable fractures and poor initial fracture reduction.^10^ Though the consequences of fracture nonunion and eventual implant failure are well-recognized, hip fracture readmission rates have largely remained unchanged from 2004 (14.3 %) to 2009 (14.5 %).^11^ Investigative studies have shown that approximately one of every six readmissions is potentially preventable with intervention, indicating a clear clinical gap.^9^ Clinicians need objective information about the quality of bone healing, which is necessary to assess their patient’s risk of catastrophic implant failure. Better methods to assess the state of bone healing post-operatively are needed to improve patient follow-up care.^9^

Radiographic assessment has long been the standard for postoperatively evaluating hip fracture union, though an accurate assessment depends on the identification of several factors such as, cortical continuity, loss of fracture line on serial radiographs, and callus size.^12-14^ Attempts have been made to standardize radiographic assessment of fractures^15-17^, but the wide variety of factors to consider leads to fracture evaluation often becoming a judgment call of the surgeon.^12^ In a comprehensive study quantifying how bone clinicians evaluated IT fractures, Chiavaras et. al found that agreement among 100 surgeons of IT fracture healing assessment was merely 0.24 (0.103–0.375) on a −1 to +1 scale of agreement, based on an intraclass correlation coefficient (ICC) score with 95 % confidence intervals. This indicates that radiography alone offers only a slight level of agreement among surgeons when assessing IT fracture healing.^12,18,19^ This method of subjective assessment comprises the cornerstone of post-operative patient care, and surgeons must balance patient mobility with the risk of implant failure. A measure of implant strain can offer an objective means of understanding the stability of a fracture site.

While the biological signs of fracture union may be difficult to track, mechanical performance of the implant and surrounding bone could be clearly visualized, providing an objective means to assess fracture stability. Initially, implant screws experience significant bending strain upon weight-bearing because the fractured bone is incapable of supporting load.^20^ As the bone heals and the fracture callus proliferates and strengthens, implant loading lessens. Clinical studies with instrumented external tibial fracture fixation healing have demonstrated the utility of measuring implant strain as an assessment tool in quantifying bone healing.^21^ A study by Richardson et al. found that when the stiffness thresholds (fracture callus stiffness was 25% of intact bone) provided a better metric for safe hardware removal than conventional analysis based on traditional timeframes, radiographs, and symptomologies. Specifically the stiffness threshold allowed hardware removal 2.3 weeks earlier than the conventional treatment control, while avoiding refractures (0 of 95 patients compared to 8 of 117 (7%) in the control).^21^ Similar results were observed by Claes et al, who also used strain measurements tracking healing.^22^ While these studies were able to assess implant strain in externally fixed devices, there is a need for user-friendly sensors to measure implant strain in patients with internal fixation during the healing process.^19^

Measuring implant loading could allow physicians to track fracture healing objectively and inform decisions regarding their patent’s treatment. For example, slowly healing patients could be prescribed altered physical therapy and activity regimes, ultrasound or electrical bone stimulation therapy, diagnostics for underlying metabolic therapies, bisphosphonates, or interventional revision before catastrophic failure.^6,7^ In addition, a clearer knowledge of fracture stability could help with communication throughout the cycle of care and inform treatment of comorbidities with therapies that could otherwise affect healing.^7^ With undiagnosed delayed fracture healing, the hardware continues to be loaded can be overloaded or fatigue and fail, requiring costly revision surgery with significant risk of morbidity and mortality.^9^ A technique to quantitatively assess IT fracture stability through standard radiography could thus improve care management and communications using existing facilities.

This study describes a novel sensor that augments sliding hip screws with an internal radiopaque indicator rod. The sensor position changes as the screw bends, screw bending to be tracked as a function of fracture loading. As the bone heals, the fracture callus will carry an increasing portion of the load, which will be reflected through less screw bending and lower sensor response. We evaluate simulated stable and unstable IT fractures and determine if implant strain detection is a viable method of assessing fracture stability using plain radiography.

**Source of Funding: This work was funded by National Institute of Health (NIH) under COBRE grant 5 P20 GM103444-07 and 1R21EB019709-01A1. (PI: Jeffrey N Anker)**

## Materials and Methods

A SHS implant was modified to enable the measurement of implant strain, specifically screw bending, across a loaded IT fracture. This strain sensing SHS (SS-SHS) screw-based implant undergoes a measurable amount of load-induced bend when a patient bears weight across an unstable IT fracture. We propose that the strain indicator can be used to interpret fracture stability using standard radiography (Figure 1). Specifically, the indicator amplifies the apparent bending of the screw during weight bearing such that it can be accurately measured from traditional radiographs. This is achieved by using a straight radio-dense tungsten indicator rod that is placed through the cannulated portion of the screw to clearly highlight bending of the screw with respect to the rod. Indicator rod movement is measured by visible separation between the bottom edge at the distal end of the sensor and the interior wall of the lag screw. With the methods below we evaluate the performance of the proposed strain sensing method in simulated stable and unstable IT fractures, to determine if implant strain detection is clinically viable.

**Figure 1:**
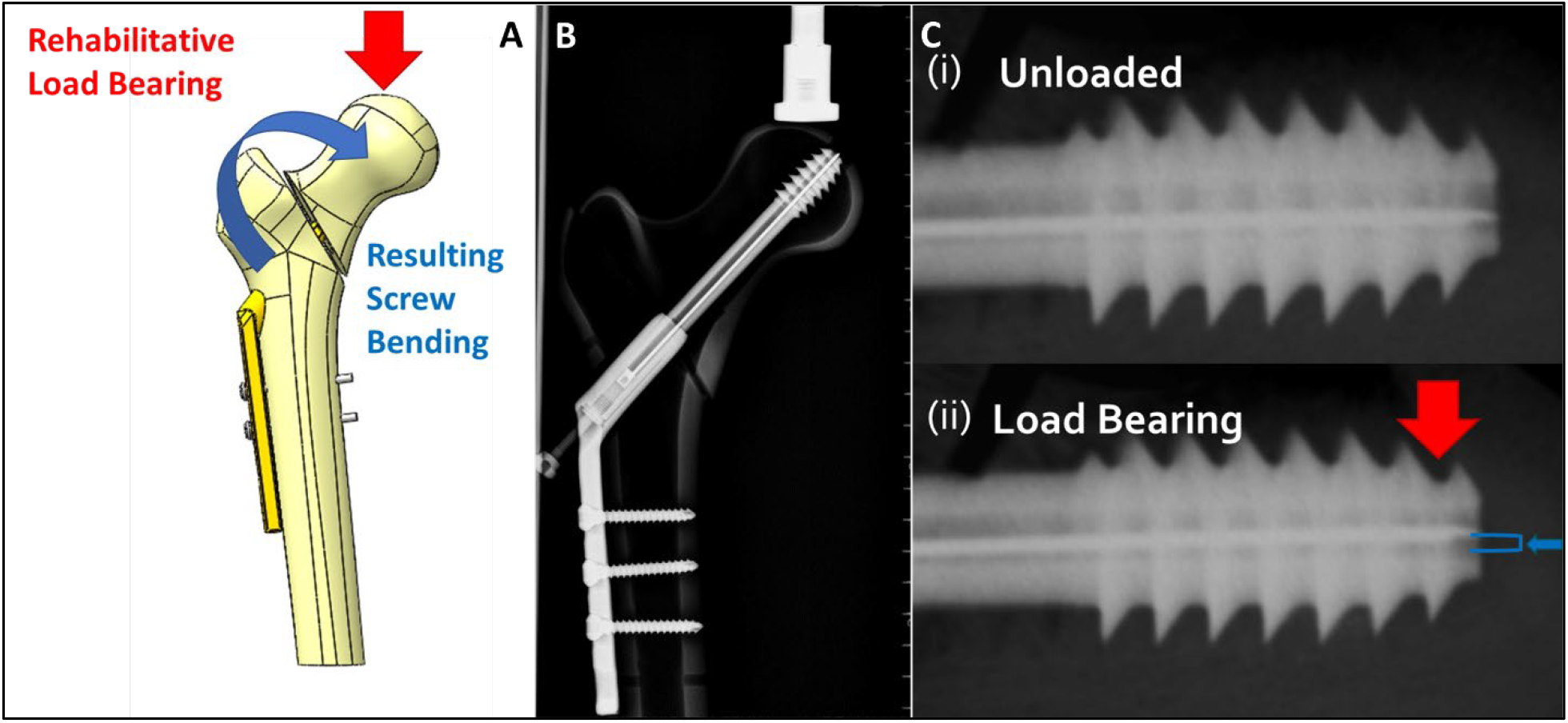
A) Computational model of a Narang sliding hip screw with sensor under a modeled loading cap from a load testing machine B) Radiograph from a Tingle TXR-325M of sliding hip screw with sensor C) Comparison of sensor position within lag screw at low (i) and high (ii) loads

A standard SHS implant (Narang Medical, New Delhi, India) was fit with the indicator rod within the 3 mm cannulated center of the lag screw. The rod itself was 0.8 mm wide, 90 mm long, and it was press fit into a 5 mm long M5 set screw base. To prevent sensor movement from the threading gap between the sensor base and the interior of the lag screw, a secondary screw secured the sensor base. Following initial computational modeling, these systems were fabricated for mechanical testing.

A computational model of an SS-SHS implant with the designed sensor was constructed and set within a computational model of a Sawbones™ (Model 3403, Sawbones, Vashon Island, Washington) medium-sized composite femur mimic with a 135° femoral neck angle, cortical and cancellous density of 1.64 g/cc and 0.27 g/cc respectively^23^, and a type A1 medial intertrochanteric fracture. The model was then imported to ANSYS, where a finite element model for proposed load testing was constructed. Tetrahedral patch conforming mesh was used to prove convergence and determine a reference solution, with a fine mesh component applied to the lag screw and sensor. Due to the focus of the study, small geometric displacements, this selection was optimized to retain geometry and maximize accuracy. All bodies had the same element size, and variables were controlled globally. To model a benchtop experiment, a loading cap was designed with a contact which permitted force transfer and slight sliding. Fracture contact was frictionless with permitted limited sliding. Mesh elements were manually selected underneath the loading cup to define contact body, and selected nodes at the bottom of the indicator rod and interior of the lag screw wall were tracked to determine sensor displacement relative to the lag screw interior. The SHS geometry was reverse engineered, and material properties for the Ti-6Al-4V titanium and tungsten were estimated to be E=104.8 GPa, v=0.31 and E=124 GPa, v=0.28, respectively (Solidworks Material Library 2019, Dassault Systèmes, Vélizy-Villacoublay, France). Force testing was applied in two one second steps with a ramp input slope of one. Each step was broken down into five sub-steps, with ten data points in total. The first step ramped from 0-1000 N, and the second ramped down to 100 N. Gravitational acceleration was applied to the entire model.

Two mechanical IT fracture models were constructed with physical Sawbones™ Model 3406 medium-sized femurs, and each was implanted with titanium SS-SHS systems (135° four-hole plates and 105 mm lag screws). One model was cut with a table saw to simulate a type 1 IT fracture, and the other was left intact to simulate a fully healed fracture. Each plate was secured with two stainless steel cortical screws and the lag screw implantation was conducted as per AO surgical guidelines with great care being taken to maintain proper tip-apex distance (>25 mm total distance from center across AP and lateral views).^24^ The lower condyles for each femur were then aligned at 12° and fixed in Fast-Cast cement (Goldenwest Inc., Cedar Ridge, CA). Each IT model was subjected to axial compression via one pre-load cycle and four loading cycles using an ESM303 axial load testing machine (Mark-10, Copiague, NY). Each loading sequence consisted of static loads from 100 N to 900 N in increments of 100 N for 20 second pauses during each load, producing 83 individual load states for each model. Radiographs were taken at each state with a Tingle TXR-325M (TXR, Cottondale, AL). Each radiograph was then analyzed (ImageJ, Version 1.51f, NIH) to track the position of the indicator rod within the lag screw.

Two cadaveric femurs of a similar size and neck angle were acquired from the Steadman- Hawkins Clinic (Patewood facility, Greenville, SC) and implanted with titanium SS-SHS systems as described in previous tests. As before, the lower condyles of both femurs were fixed at 12° in Fast-Cast cement. Specimen 1 was severed completely at far medial trochanter with a surgical saw to simulate a type A1 intertrochanteric fracture, and Specimen 2 was left intact, representing a fully healed fracture. Each femur was then mounted to an ESM303 axial load testing machine and the femoral head was subjected to a compressive loading sequence of static loads from 100 N to 900 N in increments of 100 N for 20 second pauses during each load. Radiographs of the femoral head and device were taken with a Quantum C-Arm (Ziehm Imaging, Nuremberg, Germany) at each force increment. If any implant strain was detected by the device on the first cycle, the load cycle was repeated four additional times, again under fluoroscopy. Following loading, analysis was conducted on each image to track the position of the tindicator rod within the lag screw.

## Results

The finite element model of the proposed device and experimental setup successfully converged after a simulated load cycle. Load testing shown in the model indicated that the sensor would move linearly upwards within the lag screw at a rate of 0.80 µm/N in response to implant strain. Figure 2 shows the simulated response to a triangle wave cyclic loading.

**Figure 2:**
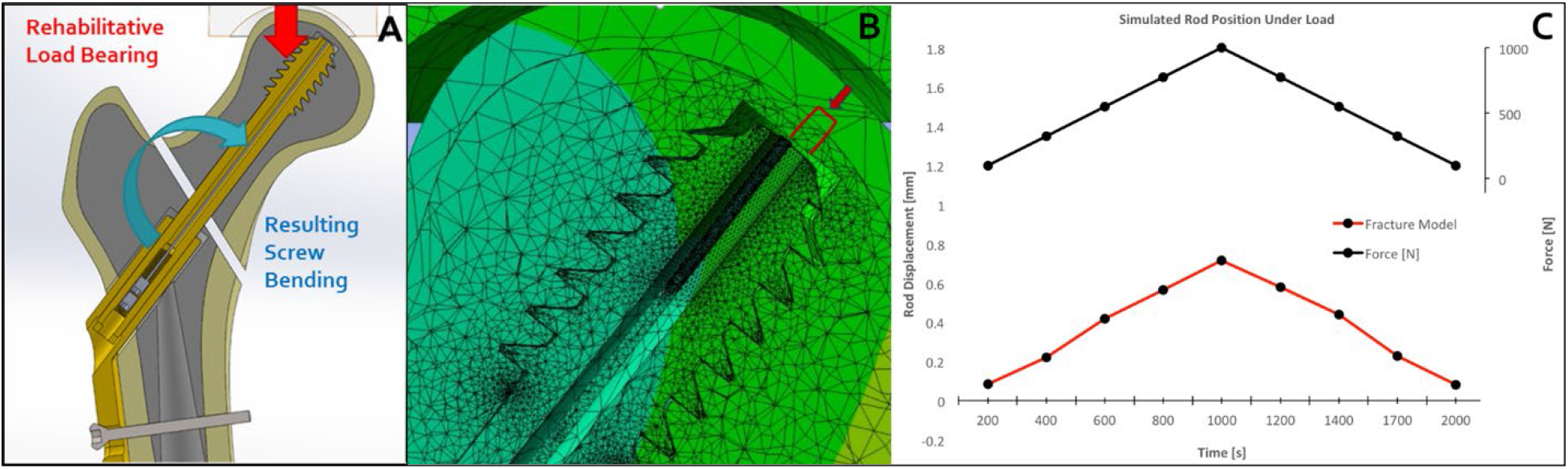
A) CAD model of Narang sliding hip screw with sensor under a modeled loading cap from a load testing machine. B) ANSYS FEA model mesh overlay colored to indicate displacement across a femoral head with an instrumented sliding hip screw implant. C) Comparison of sensor position within lag screw to applied loading curve.

Across all testing, the smallest measurable sensor movement observed via radiograph was 0.01 mm, establishing a reasonable estimate of sensor resolution. In the mechanically equivalent femur model (Figure 3), the indicator rod was clearly visible within the lag screw in each radiograph, allowing the rod base position to be determined with image analysis to within a displacement accuracy of ± 0.01 mm. In the fractured femur, the rod showed clear displacement with each change in load, starting at 0.37 ± 0.01 mm at 100 N, and continuing upwards until reaching 1.13 ± 0.02 mm at 900 N. The maximum hysteresis was 160 N with overall sensor response slope correlating to 1.0 µm/N. Due to the secure interface between the sensor and lag screw, it is believed that the observed hysteresis can be attributed to the extended dry contact between fracture pieces rather than any interface between the sensor and screw. The sensor displacement noise level was ~2.2% of displacement at 900 N. In the unfractured model, sensor motion was measurable up to 900 N.

**Figure 3:**
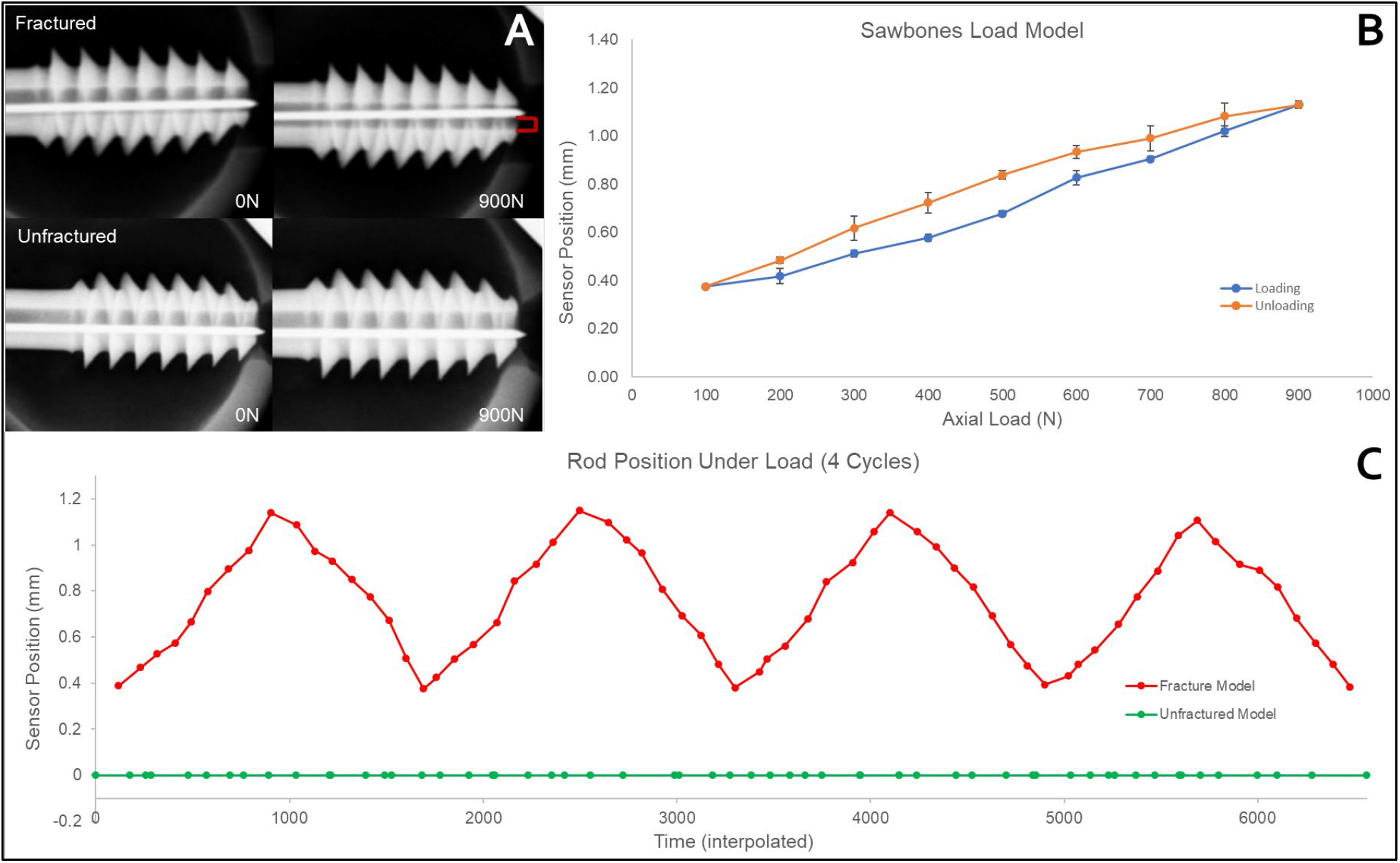
A) Loaded and unloaded radiographs of instrumented SHS lag screws within fractured and unfractured Sawbones. B) Hysteresis curve for the average across the four loading cycles, exhibiting a maximum hysteresis of 0.16 mm and a precision of ± 24.6 N. C) Time-interpolated data series charting sensor position alongside force applied to each respective model throughout the four loading cycles.

In the unfractured model, the rod exhibited no discernable movement throughout the entire load cycle. This lack of movement indicates that the screw was not exhibiting load-induced strains greater than 0.01 mm, and it can be reasonably concluded that the intact bone was significantly shielding the implant from load.

In the cadaveric femurs (Figure 4), the indicator rod was clearly visible within the lag screw in each radiograph, allowing the rod position to be determined with image analysis to within a displacement of ± 0.01 mm. In the fractured femur, the rod showed clear displacement with each change in load, starting at 0.38 ± 0.03 mm at 100 N, and continuing upwards until reaching 0.72 ± 0.06 mm at 900 N. The maximum hysteresis was 45 N with overall sensor response slope correlating to 0.80 µm/N. The sensor displacement noise level was ~7% of displacement at 900 N. In the unfractured femur, there was no discernable sensor movement throughout the entire load cycle of up to 900 N. This indicates that the screw was not undergoing load-induced strains of greater than 0.01 mm, and it can be reasonably concluded that the intact bone was significantly shielding the implant from load.

**Figure 4:**
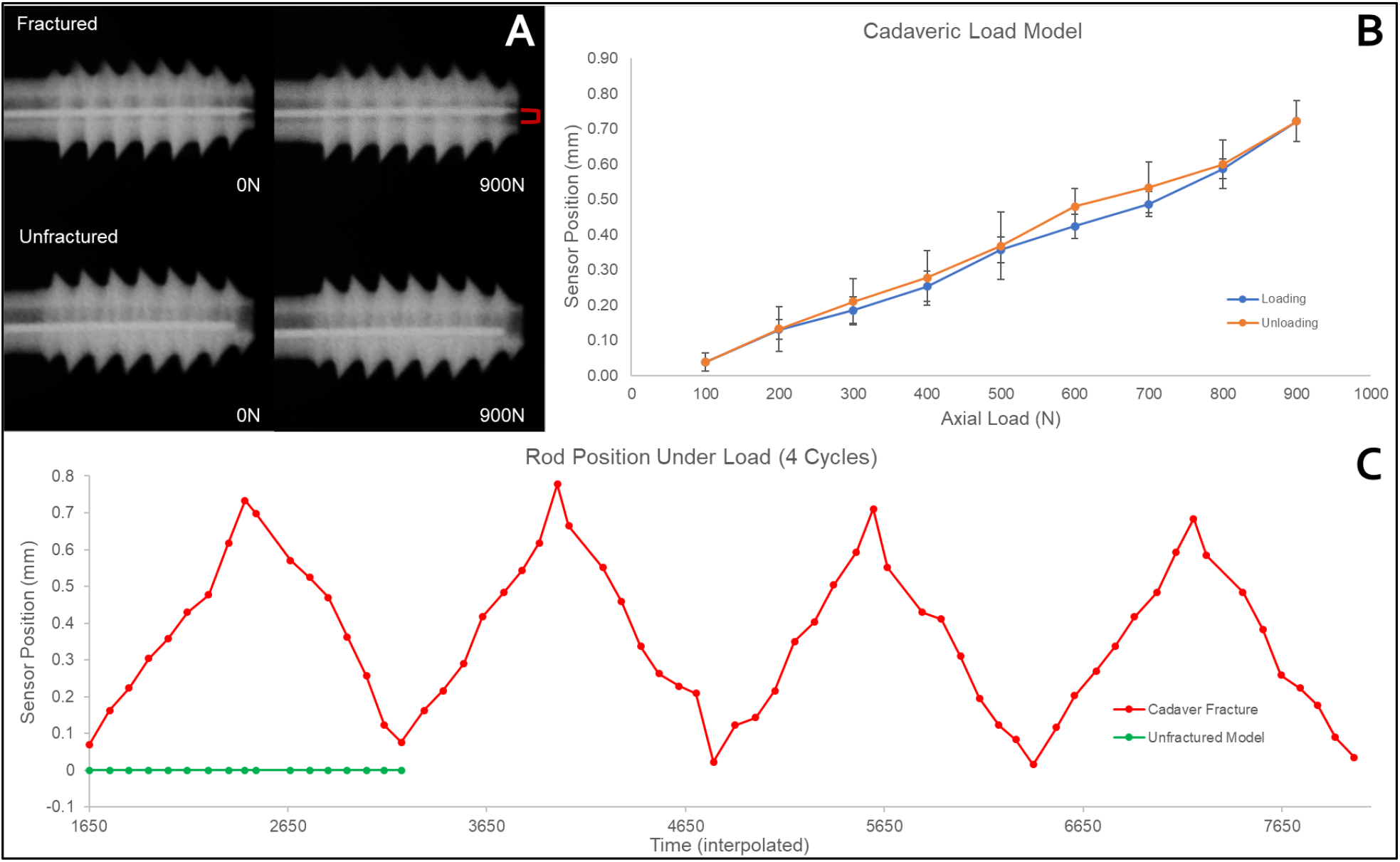
A) Loaded and unloaded radiographs of instrumented SHS lag screws within fractured and unfractured cadaver models. B) Hysteresis curve for the average across the four loading cycles, exhibiting a maximum hysteresis of 0.06 mm and a precision of ± 63 N. C) Time-interpolated data series charting sensor position alongside force applied to each respective model throughout the four loading cycles.

## Discussion

From an orthopedic perspective, fracture healing is defined in terms of restoration of a bone’s biomechanical function, especially stiffness.^25^ Implant strain sensing allows this stiffness to be measured to track healing. Moreover, with additional fatigue analysis, it could allow for informed decisions regarding post-operative activity level. Sommers et al. found that for the most common failure (screw cutout from the femur head), the number of cycles to failure in identical composite femur specimens decreased exponentially from 34,000 cycles under 800 N cyclic screw loading to only 10 cycles under 1,400 N.^10^ For context, a typical healthy 80 kg person would exert 784 N axial load across their hip during 1 leg standing and ~1,600 N axial load when walking normally with no assistance.^26^ Quality fracture reduction and fracture healing can partially shield the lag screw from such forces, but, if a patient takes enough steps on an unstable fracture that is not healing, either the hardware or the bone will eventually fatigue and fail with number of steps to failure.

When implant strain becomes visible, implant fatigue models such as Sommers et al.’s can be considered when making clinical observations (Figure 5). This in turn can inform when a patient is suitable for greater weight bearing and help guide other risk-mitigating actions later in the healing cycle. Using previously established implant characteristics and postoperative data as a starting point, the results of this work can inform the development of a simple protocol that uses observed sensor data to help guide postoperative weight bearing. For the early and most critical stages of post-operative activity (90 days), patients typically load their hips an average of 10,000 times.^27-29^ After accounting for sensor precision in our cadaveric model (± 0.06 mm/63 N) and patient bone variability, we can postulate that if patient activity does not regularly induce more than 800 N of load through the implant, measured as less than 1.50 mm of sensor movement, then the implant is at minimal risk of failure for the first month of post-operative recovery. This risk increases as loading exceeds 800 N, becoming unsustainable for our target of 10,000 cycles. After the first month, fracture healing should begin to shield the implant from dangerous loads, allowing patients to put greater force across their hip without exceeding our implant force limit of 800 N. If sensor behavior remains unchanged throughout the post-operative recovery period, then clinicians can begin taking preventative action, such as prescribing FORTEO or planning a revision surgery before total failure.

**Figure 5:**
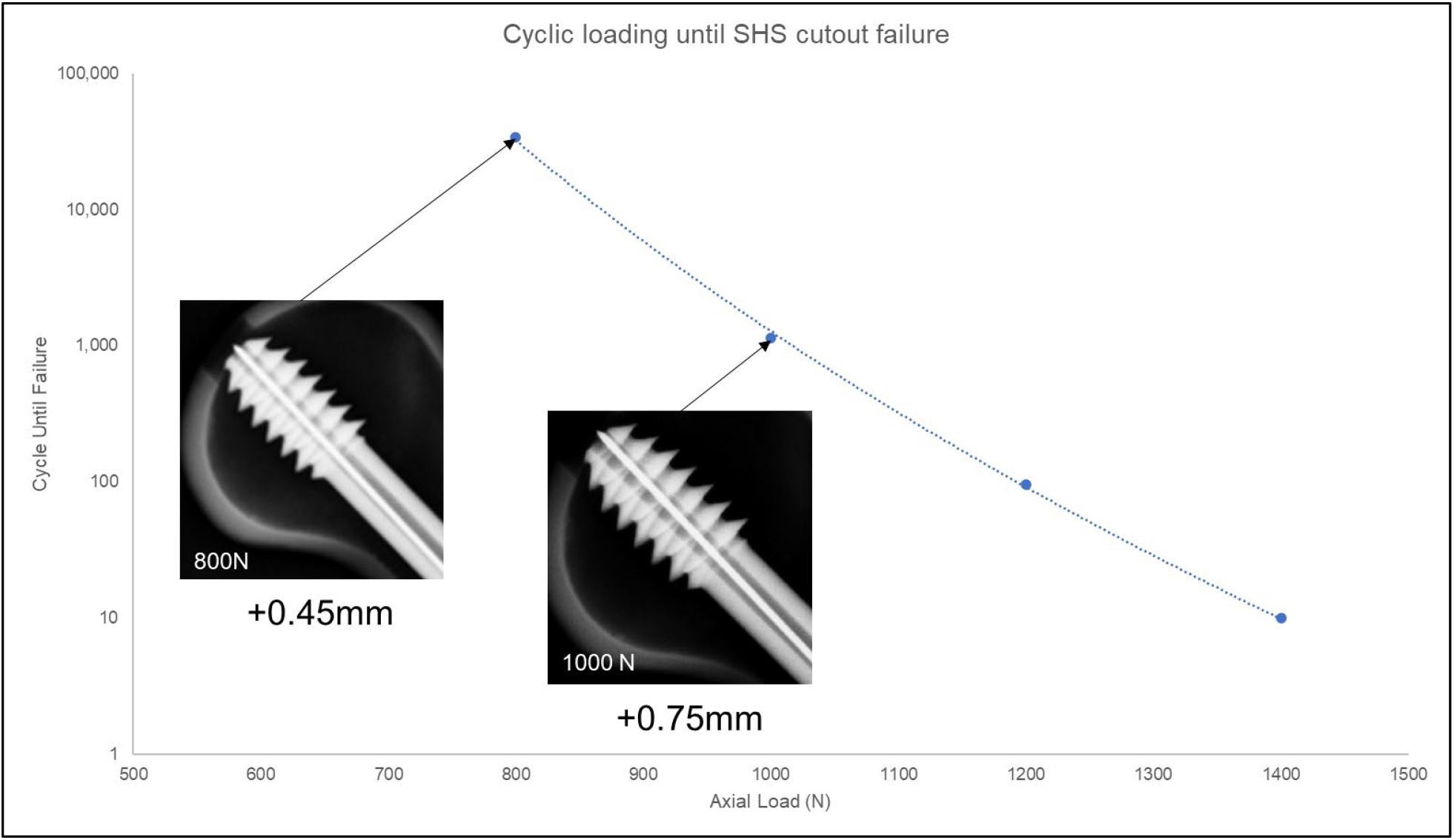
Change in sensor position can be paired with SHS predictive cutout values to aid in clinical assessment of implant longevity. Example radiographs are shown above at 600N and 100N, along with corresponding positional changes of the sensor.

Ideally, as the bone heals and the fracture callus stiffens, the load on the implant decreases during standing or walking, increasing the cycles to failure in a “race to heal.” Several clinical studies with externally fixed tibia have found that when the bone takes 80-90% of the load (leaving 10-20% of the load on the implant) hardware can be safely removed with no risk of refracture.^22^ While the hip anatomy is different, it stands to reason that at these thresholds (or possibly earlier), normal activity could be resumed with hardware retained. We find similar a result for 10^6^ cycles (about a year of walking) assuming the cutout fatigue described in Fig. 5. Thus, if the sensor can reliably detect 1/5^th^ to 1/10^th^ of the initial bending under body weight for an unstable fracture, it should be sufficient for determining safe healing thresholds. For IT fracture treatment, this means that the force precision should be ~80 N or better under an 80 kg load, which would be equivalent to ~80 µm sensor displacement.

The finite element model, composite femur, and cadaveric specimen all showed linear displacement with load (with some hysteresis especially in the composite model) and gave similar results for effective stiffness in the unstable fracture (0.8 µm/N, 1.0 µm/N, and 0.8 µm/N, respectively). Additionally, we did not observe screw bending in all intact femurs with no discernable sensor movement. We observed a 63 N maximum noise level in the cadaveric specimen, which meets our defined precision requirements. This suggests that it is possible to sufficiently track fractures as they are healing through SS-SHSs until there is minimal risk of cutout or failure during normal use.

While initial findings are promising, there are several limitations which will guide future work. First, tests were restricted to 105 mm lag screws. Since bending deflection scales with the free length of the screw cubed, shorter screws are expected to exhibit smaller deflections and an additional mechanical gain mechanism may need to be added to the indicator rod to increase sensitivity. Second, as a diagnostic aid, the effectiveness of the sensor is dependant upon how clearly an operator can read and interperate sensor position. With the current sensor design, it is crucial that it is aligned in plane with patient hip loading and that sensor position can be clearly seen in a radiograph. Future design improvements, such as integrated screw markings and an incorperated mechanical gain mechanism could greatly improve effectiveness and user experience. Third, the current cutout fatigue analysis is relatively crude, and could be enhanced with more patient specific information. Further studies can explore proper safe loading thresholds while accounting for patient body weight, activity level, and number of weeks removed from surgery.

## Conclusion

In summary, we have developed a simple tool that can quantify load sharing between a screw and developing fracture callus by making screw bending visible under radiography. As the femur is axially loaded, linear indicator rod movement corresponds with fracture instability and little to no rod movement indicates full callus formation. Quantifying such load sharing is crucial to better understanding when a fracture is healing and can allow for more informed post-operative guidance.

## Data Availability

The data that support the findings of this study are available from the corresponding author, NTC, upon reasonable request.

